# Primary Subclavian Venous Thrombosis - Multidisciplinary Collaboration Improves Early Identification and Frequency of Surgical Intervention

**DOI:** 10.1101/2025.05.04.25326953

**Authors:** Fredrik Sartipy, Klara Milles Schalling, Christian Smedberg, Victor Mill, Peter Gillgren, Jonas Malmstedt

## Abstract

Primary subclavian venous thrombosis (PSVT) is a rare but potentially disabling condition caused by chronic compression of the subclavian vein at the thoracic outlet, typically affecting young individuals. Delayed diagnosis often leads to post-thrombotic syndrome (PTS) due to missed opportunities for early surgical intervention. In Stockholm, Sweden, a regional awareness and referral program initiated in 2013 significantly increased early detection and intervention rates. Between 2013 and 2024, 283 PSVT cases were identified, with incidence rising from 0.6 to 1.5 per 100,000/year, attributed to improved detection. Early surgical intervention rose from 66% to 83%, while PTS cases requiring delayed surgery dropped by 50%. These results underscore the benefit of structured regional programs.

## Research Letter

Primary subclavian venous thrombosis (PSVT) is a rare condition that can lead to post thrombotic syndrome (PTS) with significant arm disability if not promptly diagnosed and managed. PSVT mostly affects young people and occurs as result of a long-standing compression mechanism of the proximal subclavian vein at the costoclavicular junction in the thoracic outlet ^1^. The infrequency and consequently unawareness of the condition often results in a delayed diagnosis. Combined surgical intervention with early catheter directed thrombolysis followed by decompressive surgery is a treatment strategy requiring highly specialized care, only available at few vascular units. Therefore, PSVT is an underdiagnosed and undertreated condition.

Several studies report varying incidence, with some suggesting that PSVT has an incidence rate of about 1 – 2 per 100 000 people per year ^2^. Patients with PSVT present at different levels of care. Since the correct initial diagnosis may be challenging, many patients are not identified in the acute phase (within two weeks) when interventional treatment with thrombolysis is more favorable and has better outcome. Once the condition has become chronic, the surgical treatment is more complex, and the complication rate is higher. Patients with a late diagnosis are consequently more likely to receive only conservative pharmacological treatment which is associated with a high risk of PTS of the arm ^3^. Therefore, an early identification and treatment strategy is of importance.

In the Stockholm region in Sweden, (2.4 million inhabitants), a program to improve awareness of PSVT, thereby promoting early referral for evaluation and treatment, was started 10 years ago. The program included educational activities, personal communication and written guidelines, all in collaboration with regional coagulation specialists, emergency physicians and general practitioners. We here report the incidence of PSVT in the Stockholm region and number of surgical interventions performed during the last 12 years.

During 2013 – 2024, 283 subjects were identified with PSVT in the Stockholm region, representing an average incidence of 1.0 per 100 0000 people per year, with an increase from 0.6 during the first 6 years to 1.5 during the recent 6 years. This increase is most likely a result of an increased detection rate and not a true increase. In total, 221 of these were selected for early intervention and were treated according to a standardized protocol including catheter directed thrombolysis followed by first rib resection through an infraclavicular approach ^4^. All patients that were accepted for surgery had significant arm symptoms and a high demand for a normal arm function. The proportion of patients who received early surgical treatment increased from 66% in the first 6 years to 83% in the recent 6 years (figure 1). In the same period, the number of patients with PTS where elective surgery was indicated was reduced by 50%.

**Figure 1.**
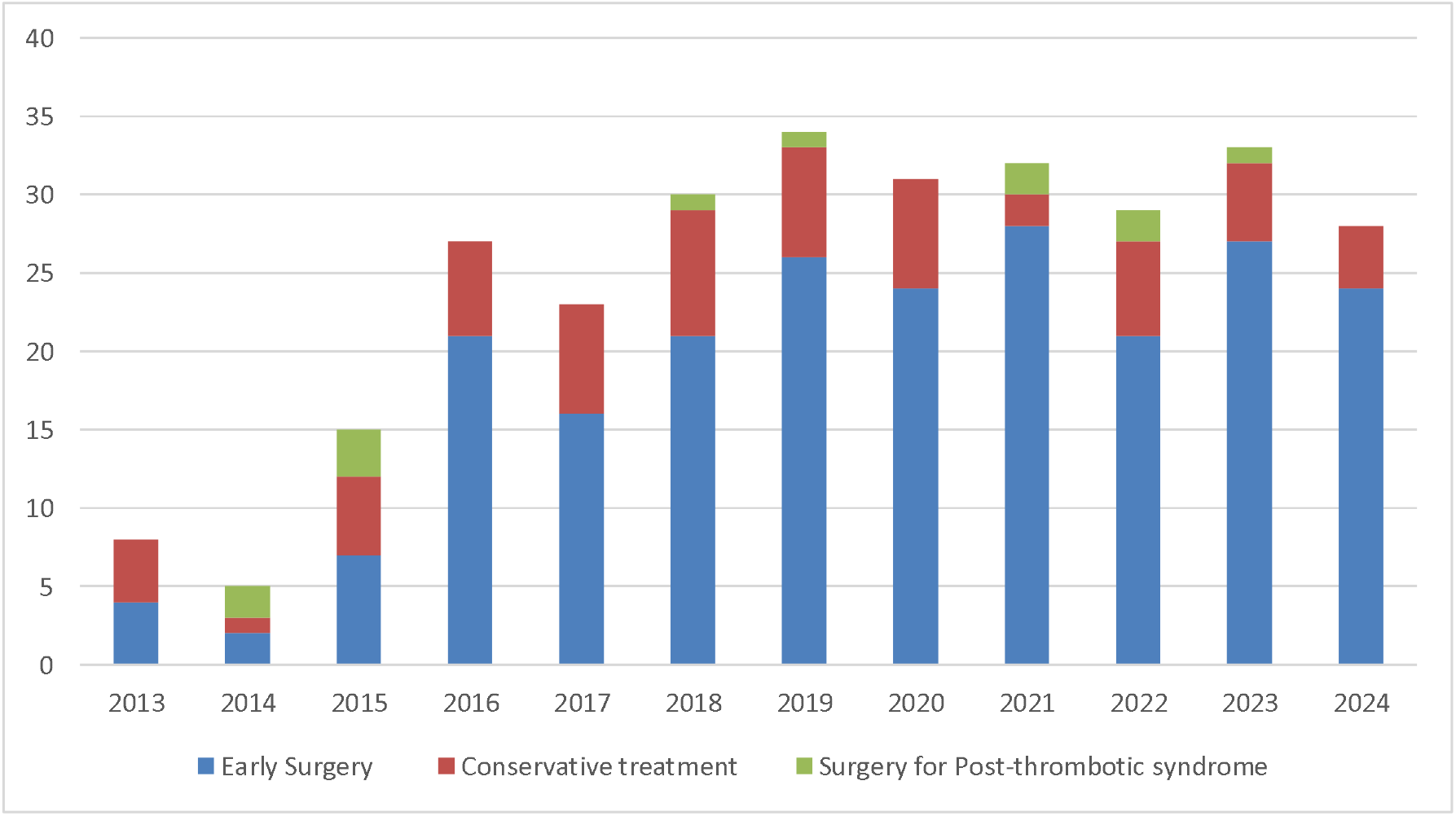
All cases of Primary Subclavian Venous Thrombosis divided by treatment modality and year 2013 – 2024 in the Stockholm region.

In comparison, in the United States, where the treatment accessibility of PSVT is relatively high, there were about 1 815 surgical procedures performed for venous thoracic outlet syndrome between 2010 – 2015 ^5^. With a population of 340 million people and an expected incidence rate of 1.0 per 100 000 people/year, an expected number of cases suitable for surgery would have been 20 400 over the same time-period, of which 9 % (corresponding to an operative incidence of 0.09/100 000 per year) were operated on either acute or chronic basis.

In conclusion, PSVT is a rare condition that requires early recognition and appropriate management to avoid the post thrombotic syndrome of the arm. Many patients are still treated conservatively with anticoagulation only and thus not evaluated for surgical intervention. This reflects the fact that surgical intervention varies significantly across different geographic areas in Sweden and worldwide.

With our strategy, we show that an active approach to this condition and establishment of a close collaboration with regional coagulation specialists and educational activities for emergency physicians and general practitioners, made it possible to improve early recognition and referral for treatment in the acute phase. This increased the turnover from conservative treatment towards an early surgical strategy, which has a more favourable outcome ^3^.

As the condition is rare, the authors advocate for further national and international education activities and centralization of treatment for PSVT.

## Data Availability

Individual participant data underlying the results reported in this Article will be made available after de-identification, alongside a data dictionary, study protocol, and informed consent form. The data will be available at Article publication and for 10 years subsequently. Data will be shared for individual participant data meta-analysis with other members of the research community who have an affiliation to a recognised medical university. Data will only be shared with investigator support and after approval of a proposal, and with a signed data access agreement. Additional restrictions apply according to Swedish law.

